# Diagnostic Accuracy and Potential Resource Savings of Pooled Sputum Testing with Xpert MTB/RIF Ultra for Tuberculosis among adults in Vietnam: A Cross-Sectional Study

**DOI:** 10.64898/2026.03.31.26349825

**Authors:** Han Thi Nguyen, Andrew James Codlin, Luan Nguyen Quang Vo, Nga Thi Thuy Nguyen, Rachel Forse, Ha Thi Minh Dang, Lan Huu Nguyen, Hoa Binh Nguyen, Luong Van Dinh, Ha Thu Doan, Hung Van Nguyen, Jacob Creswell, Tushar Garg, Ana Isabel Cubas Atienzar, Rachel Louise Byrne, Vibol Iem, Bertie Squire, Lina Davies Forsman, Tom Wingfield, the Start4All Vietnam Team

## Abstract

**Objectives:** A pooled testing algorithm for tuberculosis (TB), in which sputum specimens from multiple individuals are tested in pools with individual testing of positive pools, can optimise diagnostic resources. This study evaluated the diagnostic accuracy and cartridge savings of pooled testing with the Xpert MTB/RIF Ultra assay (Xpert-Ultra) relative to individual Xpert-Ultra testing.

**Methods:** We conducted a cross-sectional study among 2,396 adults (≥15 years) with presumptive TB enrolled between July 2024 and February 2025, through facility-based case finding (FBCF) and community-based case finding (CBCF). Participants submitted two sputum specimens. The first underwent individual Xpert-Ultra testing; remnant specimens were combined into four-specimen pools and tested again with Xpert-Ultra. The second specimen was used to inoculate liquid culture (BACTEC MGIT). Data were used to simulate an up-front pooled testing strategy; sensitivity and specificity of this approach was estimated against culture, and cartridge use was compared with individual Xpert-Ultra testing.

**Results:** Of 2,396 participants, 395 (16.5%) had a positive Xpert-Ultra and/or culture, including 360/912 (39.5%) in FBCF and 35/1484 (2.4%) in CBCF. The pooled testing approach had sensitivity of 82.4% (95% confidence interval [CI], 77.9-86.3) and specificity of 98.5% (97.8-99.0) compared to culture, with lower sensitivity than individual Xpert-Ultra testing (86.5%, 82.4-89.9) but high specificity (98.1%, 97.4-98.7). Sensitivity of pooled testing was lower in CBCF (59.1%, 36.4–79.3) than in FBCF (84.0%, 79.5–87), whereas cartridge savings were greater in CBCF (69.1% vs 9.6%). The pooling strategy reduced Xpert-Ultra cartridge use by 46.5%, saving USD 14,447.

**Conclusions:** Pooled Xpert-Ultra testing among adults appears resource-efficient for TB screening in Vietnam. As sensitivity is lower compared to individual Xpert-Ultra testing, particularly for paucibacillary disease, these losses should be carefully weighed against gains in affordability and expand access to molecular testing. Careful, context-specific implementation is essential to maximise programmatic benefit while minimising missed persons with TB.

## Introduction

Tuberculosis (TB) remains a significant global health threat. In 2024, 10.7 million people developed TB, and 1.2 million died (1). Vietnam is among the highest TB burden countries, with an estimated incidence rate of 182 per 100,000 population (1).

The Xpert MTB/RIF Ultra (Xpert-Ultra) assay is a World Health Organization (WHO) -recommended rapid molecular test for TB (2) with superior performance to smear microscopy (3). However, its use in screening campaigns, particularly in congregate (4), decentralised or community settings (5), and low-incidence settings (6), remains limited due to cost (7), tightening global funding for TB programmes and operational constraints limiting large-scale deployment, including infrastructure and workforce requirements (8). Pooled testing, whereby sputum specimens from multiple individuals are combined and tested as a single specimen, has been proposed to improve testing efficiency (9-11) and reduce costs (12, 13). This testing strategy has been successfully used in infectious disease programmes, particularly during the COVID-19 pandemic (15) and to support TB service continuity (16). Abdurrahman et al. (17) showed that three- and four-specimen pooled testing using the Xpert MTB/RIF (Xpert) assay retained high positive percent agreement (PPA) with individual Xpert testing, while significantly reducing cartridge consumption during an evaluation in Nigeria. Iem et al. (18) reported similar performance and cost savings using pooled testing during community-based case finding (CBCF) in Laos. In light of this growing body of evidence, pooled testing was recently endorsed by the WHO to improve efficiency in TB diagnosis (14).

Adoption of pooled testing in high-burden settings such as Vietnam remains limited. The country’s National TB Control Programme (NTP) primarily relies on facility-based case finding (FBCF), with CBCF as a complementary strategy to reach underserved populations. Therefore, evidence on the real-world implementation of pooled Xpert-Ultra testing is needed to inform national policy and guide scale-up decisions.

## Methods

### Design and aim

We conducted a cross-sectional, multi-site study to compare the diagnostic accuracy and cartridge savings of a theoretical upfront pooled testing strategy using four-specimen pools versus individual Xpert-Ultra testing between July 2024 and February 2025. Four-specimen pools were selected based on prior evidence demonstrating acceptable sensitivity and efficiency (5). Sample size followed the multi-country study protocol, which targeted approximately 600 participants per site; in Vietnam, four sites were included (two FBCF and two CBCF sites), yielding a total sample size of 2,396 participants.

### Setting

Study recruitment occurred in Hanoi and Ho Chi Minh City (HCMC), Vietnam’s most populous provinces, with a combined population of approximately 20 million and more than 25,000 TB treatment notifications reported in 2024 (19). FBCF was conducted at the Vietnam National Lung Hospital (VNLH) in Hanoi and Pham Ngoc Thach Hospital (PNTH) in HCMC. CBCF events were organised across Hanoi (Ba Dinh, Hoan Kiem, Hoang Mai) and HCMC (Districts 06, 08, Binh Chanh) (Figure 1). All samples collected in Hanoi were processed at the VNLH, and all samples collected in HCMC were processed at PNTH.

### Participant recruitment

During FBCF, individuals aged ≥15 years presenting with a cough of any duration were eligible for recruitment. Twenty CBCF events were conducted (10 in each province), where individuals aged ≥15 years were similarly eligible for enrolment. Community members were mobilised by government health staff and local organisations (20, 21). At CBCF, participants underwent verbal WHO-aligned four-symptom screening and chest X-ray (CXR) in accordance with Ministry of Health guidelines (22). CBCF participants were eligible for enrolment if they met any of the following criteria: 1) cough of any duration, fever, weight loss and/or night sweats, 2) abnormal CXR (radiologist and/or qXR computer-aided detection software, Qure.ai, India, threshold≥0.3), 3) contact with a person who had pulmonary TB, and/or 4) self-reported HIV infection. Exclusion criteria included receipt of TB treatment within 60 days, inability to produce sputum, and/or severe illness. All participants provided written informed consent.

### Sample Collection and Testing

Each participant provided two sputum specimens, collected at least 30 minutes apart. The first specimen was individually tested using the Xpert-Ultra according to manufacturer instructions. The remaining material from the first sputum specimens wa then pooled in groups of four (0.5 mL each; total 2 mL) and tested using Xpert-Ultra (Supplementary Figure S1). The second specimen underwent liquid culture (MGIT 960, BACTEC, BD Diagnostics, USA) following standard procedures (23).

### Statistical analysis

For descriptive analyses of participant recruitment and testing (Tables 1, 2 and 3), we constructed an expanded microbiological reference standard (eMRS) to classify participants as having TB or not having TB. Participants were defined as TB-positive if they had at least one positive result from individual Xpert-Ultra and/or positive culture. Participants were defined as TB-negative if they had two non-positive results from individual Xpert-Ultra and culture testing. This eMRS was not used to assess the diagnostic accuracy of pooled vs individual Xpert-Ultra testing (Table 4); instead, MGIT culture results were used as the reference standard, in line with World Health Organization guidance on TB diagnostic assay evaluations (2).

Participant characteristics were summarised and compared by setting (FBCF vs CBCF) and province (Hanoi vs HCMC), using the chi-square and Mann-Whitney U tests. Pooled test results were summarised by the number of individually positive specimens per pool. Among participants with TB according to the eMRS, pooled semi-quantitative grades were compared with their paired individual test grades. Diagnostic accuracy for a simulated upfront pooled testing strategy, whereby only individuals with positive pooled test results were eligible for follow-on individual testing, was estimated using culture as the reference standard (excluding NTM/contaminated cultures) and compared against the sensitivity and specificity of an individual testing strategy, using generalised estimating equations with robust standard errors, accounting for clustering by participant and stratified by setting and province (24).

We estimated cartridge use under an upfront pooled Xpert-Ultra testing approach relative to individual testing. Costs were calculated using the local cartridge price (VND 340,200 per cartridge as of 13/8/2025, Nam Phuong Technique Company Limited; Hanoi, Vietnam; equivalent to USD 12.98 at an exchange rate of USD 1=VND 26,210 per www.xe.com).

All analyses were conducted using Stata version 15 (StataCorp, College Station, USA). Statistical tests were two-tailed, and statistical significance was defined as a p<0.05.

## Results

Among the 2,396 participants enrolled, 912 (38.1%) were from FBCF and 1,484 (61.9%) from CBCF. Overall, 395 (16.5%) were classified as having TB according to the eMRS, with prevalence of 39.5% in FBCF and 2.4% in CBCF (360/912 vs 35/1,484; p<0.001). The median age was 55 years (interquartile range [IQR] 39.5-66), and 51.9% of participants were male (Table 1).

Of the 599 pooled tests performed, 171 (28.5%) had positive Xpert-Ultra results (Table 2). Detection rates increased with the number of positive component specimens: 77.9% (74/95) for pools containing one positive component specimen, 95.9% (47/49) for two, 100% for three (36/36) and four (12/12). Among the 407 pools composed entirely of component specimens that were individually negative on Xpert-Ultra, two (0.5%) produced positive pooled Xpert-Ultra results. Both were from CBCF participants in HCMC and included culture-positive specimens.

Among 395 participants with TB according to the eMRS, pooled Xpert-Ultra test results were negative in 45 (11.4%) (Table 3). Of the 46 participants with culture-positive but individual Xpert-Ultra-negative results, 20 (43.5%) would never have been individually tested in an upfront pooled testing strategy because their pooled test result was negative, while the remaining 26 (56.5%) would have been individually tested and had their diagnosis missed during individual testing. Similarly, among the 49 participants with Trace individual Xpert-Ultra results, pooled test results were negative for 18 (36.7%), resulting in exclusion from follow-on testing (and missed diagnosis) if an upfront pooled testing strategy had been employed. Positive concordance between individual and pooled test results was high for higher semi-quantitative grades. Among participants with Very Low and Low individual results, pooled test results were positive in 36/37 (97.3%) and 129/135 (95.6%), respectively. All participants with Medium (57/57, 100%) and High (71/71, 100%) individual grades had positive pooled test results.

A simulated upfront pooled Xpert-Ultra testing strategy had a sensitivity of 82.4% (95% confidence interval [CI]: 77.9-86.3%) and specificity of 98.5% (97.8-99.0%), compared with culture (Table 4). Sensitivity was significantly lower than individual Xpert-Ultra testing (86.5% [82.4-89.9%]; p<0.001), (difference of 4.1%), while specificity was significantly higher (98.1% [97.4-98.7%]; p=0.008) (difference of 0.4%). In FBCF, pooled testing had a sensitivity of 84.0% (79.5-87.8) compared with 87.7% (83.6-91.1) for individual testing (p=0.001). During CBCF, sensitivity was 59.1% (36.4-79.3) for pooled testing and 68.2% (45.1-86.1) for individual testing (p=0.167). By province, pooled testing showed a sensitivity of 70.1% (59.0-80.6) in Hanoi and 85.7% (80.8-89.6) in HCMC, compared with 81.3% (70.7-89.4) and 87.9% (83.4-91.6), respectively, for individual testing (p=0.005 for Hanoi; p=0.014 for HCMC). Specificity remained high across provinces for both approaches. The distribution of semi-quantitative Xpert-Ultra grades by province is shown in Supplementary Table S1. Compared with HCMC, Hanoi had a higher proportion of culture-positive cases with negative pooled Xpert-Ultra results (26.3% vs 6.7%) and fewer medium/high grades.

A simulated upfront pooled Xpert-Ultra testing approach would have reduced cartridge use by 46.5%, from 2,396 to 1,283, yielding an estimated cost saving of USD 14,447, equivalent to USD 6.03 per participant. Reductions in cartridge use and associated costs were more pronounced during CBCF (69.1%) compared to FBCF (9.6%). In FBCF, pooling was associated with cost savings in Hanoi (−34.7%; USD 4.50 per participant) but increased costs in HCMC (+15.4%; USD 1.99 per participant). In contrast, substantial savings were observed in CBCF in both Hanoi (−69.7%; USD 9.05 per participant) and HCMC (−68.4%; USD 8.87 per participant) (Table 5).

## Discussion

This cross-sectional diagnostic accuracy study evaluated the performance of four-specimen pooled Xpert-Ultra testing versus individual Xpert-Ultra testing in both community- and facility-based settings. Our findings indicate that pooled testing can substantially reduce cartridge use, but at the expense of sensitivity, particularly in paucibacillary disease.

The observed sensitivity of 82.4% for a four-specimen upfront pooled testing strategy relative to culture is comparable to previous studies from Laos (85.7% with four-specimen pools) and Nigeria (86.3% with three- and four-specimen pools); however, those studies reported PPA with Xpert rather than sensitivity against culture (17, 18). Reduced detection among trace-positive results is typically associated with low bacillary burden in asymptomatic or pauci-symptomatic individuals, children, people living with HIV, or after recent TB treatment (<5 years) (25).

Whereas the study in Nigeria (17) evaluated pooled testing exclusively in FBCF and the study in Laos (18) focused on CBCF campaigns, our study includes both screening settings. This allowed assessment of pooled Xpert-Ultra performance across heterogeneous TB screening populations, highlighting that pooling may not be appropriate in all settings.

Pooled Xpert-Ultra showed higher sensitivity in FBCF than in CBCF. However, efficiency gains from pooling were greater in CBCF, where TB prevalence was lower. Sensitivity varied by province, with lower pooled sensitivity observed in Hanoi than in HCMC, consistent with geographic variation in TB burden in Vietnam (26). This difference appeared to be driven primarily by variation in bacillary burden, reflecting differences in the populations screened, as a higher proportion of lower semi-quantitative grades in Hanoi may increase the likelihood of false-negative pooled test results due to dilution (Supplementary Table S1). These findings suggest that bacillary burden, in addition to TB prevalence, should inform implementation of pooled testing. Pooled testing performed well among individuals with higher bacillary burden, with detection rates of 95.6% for low-grade and 100% for medium and high grades, consistent with prior evidence (27). In contrast, sensitivity declined among participants with trace-positive results and those who were culture-positive but Xpert-Ultra-negative, reflecting the known challenges of detecting paucibacillary disease across diagnostic approaches. The same challenge has been reported with the evaluation of oral and sputum swab, near-point of care molecular assays(28). This supports targeted follow-up or repeat screening after negative pooled results, particularly among individuals with abnormal CXR findings.

In FBCF, higher TB prevalence and more advanced disease increase both bacillary burden and the likelihood that pools contain more than one individually positive specimen, thereby partially offsetting the effects of dilution inherent to pooled testing. In contrast, CBCF typically identifies earlier-stage, and often paucibacillary TB. Nearly all positive pools from CBCF contained only a single positive specimen, increasing the risk of false-negative pooled test results due to dilution (8,9). In such settings, complementary strategies, including CXR-based triage or repeat testing among high-risk individuals, may help mitigate missed diagnoses.

Beyond diagnostic performance, pooled testing can offer efficiency gains. Our modelling analysis estimated a 46.5% reduction in cartridge use overall, with greater reductions in CBCF (69.1%) than in facility-based screening (9.6%). However, efficiency varied substantially by province within facility-based settings. These findings suggest that pooled testing may be best prioritised in high-volume, low-prevalence settings, such as CBCF campaigns, where a large number of individuals were screened with low TB prevalence, compared to more targeted testing in FBCF. In FBCF, pooling remained cost-saving in Hanoi but was associated with increased costs in HCMC, indicating that pooled testing may be less efficient in higher-burden settings with a greater proportion of positive pools. Our results are consistent with Abdurrahman et al. (17), who reported a 31% cartridge reduction in Nigeria, and Iem et al. (18), who demonstrated a 46% reduction in cartridge use during CBCF campaigns in Laos. These findings highlight that, beyond overall TB prevalence, the distribution of positive pools and underlying bacillary burden are key determinants of pooled testing efficiency. Overall, pooled testing remains economically attractive in resource-limited settings, particularly in low-prevalence contexts.

Efficiency gains could be further optimised by leveraging chest radiography and AI-generated risk scores to triage individuals to individual or pooled Xpert-Ultra testing. A modelling study showed greater cartridge savings than universal upfront testing through risk stratification, concentrating pooled testing in lower-risk groups (29). This strategy may be particularly relevant in higher-burden facility-based settings, where indiscriminate pooled testing may be inefficient. It may also extend pooled testing to higher-prevalence settings; however, real-world performance and feasibility warrant prospective evaluation (29). Additional strategies, including repeat or targeted re-screening of high-risk groups and adjunctive testing (eg, urine LAM in people living with HIV) (29), may also help mitigate missed cases in CBCF settings (30). Similar challenges occur with other approaches for paucibacillary disease, including oral swab-based testing, suggesting no single strategy fully addresses this limitation (28).

Our findings highlight the need to balance diagnostic sensitivity against efficiency gains. At an individual level, pooled testing is less sensitive, particularly for people with paucibacillary disease. At a programme level, its value depends on whether efficiency gains are used to expand access to molecular testing. If pooled testing replaces, rather than expands, existing testing capacity, reduced sensitivity may result in fewer individuals being diagnosed with TB. Effective programmatic implementation requires careful integration within diagnostic algorithms, including clinical assessment, CXR-based triage, and follow-up testing after negative pooled results. Linkage to care and treatment follow-up should be ensured. Acceptability and ethical implications of reduced sensitivity warrant further study.

Strengths of this study include its large sample, the inclusion of both community- and facility-based screening, and the use of culture as the reference standard. Several limitations should be acknowledged. First, children and other priority populations like PLHIV were not included, limiting generalisability to specific populations. Given the reduced sensitivity of pooled Xpert-Ultra testing for paucibacillary disease and the challenges of specimen collection in children, caution is warranted when considering pooled testing in this vulnerable group. Second, the study population was subject to selection bias, with a high proportion of TB-positive individuals recruited from high-volume, urban health facilities who are likely to have been pre-screened for TB at lower-level healthcare facilities and referred. Third, facility-based screening primarily included symptomatic individuals with cough, which may have contributed to higher detection rates compared with lower-level facilities or routine programmatic testing. Consequently, the performance of pooled testing observed here may not be generalisable to other facility-based settings with lower TB prevalence or milder disease profiles. Fourth, the economic analysis was limited to cartridge savings and did not account for broader health system costs or consequences of missed diagnoses, underscoring the need for future cost-effectiveness evaluations under routine programmatic conditions.

## Conclusion

Upfront pooled Xpert-Ultra testing can improve programmatic efficiency by reducing cartridge use and costs, but lowers sensitivity, particularly for people with paucibacillary disease. Sensitivity losses were most pronounced during CBCF, whereas efficiency gains were limited in FBCF with higher TB prevalence. These findings highlight the importance of careful, context-specific implementation, supported by risk stratification, to balance efficiency gains against missed diagnoses. Pooled testing strategies should be integrated with complementary approaches, including repeat or targeted re-screening of high-risk groups and adjunctive testing. To achieve population-level benefit, efficiency gains from pooled testing should be used to expand screening coverage rather than replace existing testing capacity.

## Data Availability

De-identified participant data were accessed and analysed within approved research collaborations. Data are not publicly available, and any further access is subject to approval by the relevant Vietnamese authorities

## Declaration

### Ethics Approval and Consent to Participate

Ethics approval was obtained from the institutional review boards of the Liverpool School of Tropical Medicine (22-084), the World Health Organisation (ERC.0003921 for the master protocol and ERC.0004084 for Vietnam protocol), the Vietnam National Lung Hospital (Approval No. 66/23/CN-HDDD-BVPTU), and Pham Ngoc Thach Hospital (Approval No. 1382/PNT-HDDD).

## Acknowledgements

We thank all study participants, doctors, nurses, and laboratory technicians at the Vietnam National Lung Hospital and Pham Ngoc Thach Hospital for their dedicated contributions. We gratefully acknowledge the support of local healthcare staff from district and commune health centres who supported CBCF events in selected districts of both provinces: Ba Dinh, Hoang Mai and Hai Ba Trung districts in Hanoi and District 06, District 08 and Binh Chanh district in HCMC. We also thank study coordinators H Pham, H Le, TNH Ong, VTC Nguyen, and TTT Tran for managing recruitment.

## Contributors

AJC, HN, LNQV, and TW conceived the study and drafted the protocol. HN and AJC led manuscript preparation and jointly conducted the analyses. HN, LNQV, and AJC oversaw enrolment and coordination. NBH, DVL, and TW, together with LNQV, supervised the work. HBN, LVD, and HTD provided resources and facilitated study implementation through the National TB Programme. TW and LNQV were responsible for funding acquisition and resource mobilisation. HN, AJC, LNQV, and LDF verified the data. All authors contributed to data interpretation, reviewed the manuscript critically for important intellectual content, and approved the final version.

